# Simultaneous Monitoring of Disease and Microbe Dynamics Through Plasma DNA Sequencing in Pediatric Patients with Acute Lymphoblastic Leukemia

**DOI:** 10.1101/2021.03.30.21253149

**Authors:** Valentin Barsan, Yuntao Xia, David Klein, Veronica Gonzalez-Pena, Sarah Youssef, Yuki Inaba, Ousman Mahmud, Sivaraman Natarajan, Vibhu Agarwal, Yakun Pang, Robert Autry, Ching-Hon Pui, Hiroto Inaba, William Evans, Charles Gawad

## Abstract

Treatment of acute lymphoblastic leukemia (ALL) necessitates continuous risk assessment of leukemic disease burden as well as infections that arise in the setting of immunosuppression and myelosuppression. This study was performed to assess the feasibility of a hybrid capture NGS panel to longitudinally measure molecular leukemic disease clearance and microbial species abundance in 20 pediatric patients with ALL throughout induction chemotherapy. This proof-of-concept helps establish a technical and conceptual framework that we anticipate will be expanded and applied to additional patients with leukemias, as well as extended to additional cancers. Molecular monitoring can help to accelerate the attainment of insights into the temporal biology of host-microbe-leukemia interactions, including how those changes correlate with and alter anticancer therapy efficacy. We also anticipate fewer invasive bone marrow aspirations or biopsies will be required as these methods improve with standardization and are validated for clinical use.

**Key Points:**

- A custom-designed hybridization capture next generation sequencing (NGS) panel noninvasively and quantitatively measures leukemic disease burden and microbial species abundance through serial peripheral blood draws throughout induction chemotherapy.
- Sequencing of circulating tumor DNA (ctDNA) alongside microbial cell free DNA (mcfDNA) in the same sample reveals the effectiveness of chemotherapy alongside the immunosuppressive consequences of cytotoxic therapy.
- Efforts to incorporate ctDNA and mcfDNA NGS monitoring in higher risk patient populations have the potential to uncover mechanisms of response, resistance, and infectious complications that correlate with patient outcomes.

## Introduction

Cell-free DNA (cfDNA) sequencing has shown promise as a noninvasive diagnostic tool for evaluating maternal and child health^1, 2^, detecting cancer earlier and monitoring treatment response^3-6^, and identifying infectious microbes with less bias^7, 8^. ALL is the most common pediatric cancer and, despite dramatic improvements in outcomes over the past fifty years, remains one of the leading causes of pediatric cancer-associated morbidity and mortality^9^. Further, due to the immunosuppressive and myelosuppressive consequences of current cytotoxic therapies, infectious complications are the major cause of treatment-associated mortality^10^.

The presence of cfDNA in plasma has been known for decades^11, 12^. Predicated upon advances in DNA sequencing technologies, circulating tumor DNA (ctDNA) is currently being evaluated as a biomarker for the early detection of malignancies, as well as a tool to monitor response and guide treatment^4-6, 13, 14^. Much of the work to-date has been performed on solid tumors where ctDNA monitoring has increasingly demonstrated diagnostic and prognostic value that has the potential to improve the outcomes of cancer patients^15-19^. Cell-free RNA sequencing and nucleosome imprinting have determined that hematopoietic cells are the largest contributors to circulating nucleic acid pools in healthy individuals^20, 21^, suggesting that ctDNA monitoring may be even more sensitive in the detection of tumors of hematopoietic origin when compared to solid tumors. Large studies of ctDNA derived from lymphomas have demonstrated improved risk assessment when used to monitor minimal residual disease^22-24^. Previous groups have applied next generation sequencing (NGS) on cellular pediatric leukemia samples to characterize mutational landscapes at diagnosis^25-28^ and relapse^29-31^, as well as measure immunoglobulin clonality^32^ as a sensitive biomarker of residual disease. Limited cfDNA sequencing studies have been performed on acute leukemias^23, 33^, and to date, we are unaware of studies that have evaluated ctDNA in pediatric leukemia patients.

The identification and quantification of non-human DNA in plasma has also shown promise as a strategy for diagnosing infectious diseases^7^ as well as monitoring the human virome^34-36^. Studies have shown that microbial cell-free DNA (mcfDNA) sequencing has similar sensitivity to standard methods for common causes of bloodstream infection, with an increased capacity to more rapidly detect rare species as well as pathogens otherwise difficult to detect through conventional cell culture methods^8, 37^. For example, shotgun mcfDNA sequencing has been recently shown to predict impending infections in pediatric cancer patients^38^. Additional improvements in the sensitivity of mcfDNA sequencing could further establish the clinical utility of microbial sequencing for diagnosing and predicting the presence of infectious complications. One strategy for increasing the sensitivity of mcfDNA detection is to enrich for microbe-specific sequences.

In the present study, we sought to develop an approach that would simultaneously monitor the two leading causes of mortality in pediatric ALL patients: 1) disease persistence or relapse and 2) infectious microbes in the setting of immunosuppression. To accomplish this, we developed and evaluated a novel capture-based ultra-deep sequencing strategy that enriched for leukemia-specific mutations and potential microbes in the plasma of ALL patients as they underwent induction chemotherapy. We found that we can detect and monitor leukemia in most patients and identified specific patterns of ctDNA dynamics as patients underwent treatment. Further, by comparing cellular to ctDNA variant allelic fractions (VAFs), we found that performing invasive bone marrow examinations at diagnosis to isolate malignant cells may be unnecessary for genomic analyses, as similar variants are detected at comparable frequencies in both sample types. Finally, we found that the human infectome is dynamic in ALL patients during the first 6 weeks of treatment, with evidence for widespread reactivation of herpes and polyoma viruses.

## Methods

### Patient Cohort, Sample Processing, and Sequencing Library Preparation

20 patients (8 females, 12 males, average age 7.9 years) enrolled on Total Therapy XVII for Newly Diagnosed Patients With Acute Lymphoblastic Leukemia and Lymphoma (NCT03117751) at St. Jude Children’s Research Hospital had serial samples collected during routine blood draws after approval of the Institutional Review Board and informed consent for research use. The protocol was reviewed and approved by the St. Jude and Stanford Institutional Review Boards. Sample workflow (Figure 1A) involved DNA extraction followed by sequencing of libraries prepared through direct ligation and capture-based enrichment using a custom panel. Bone marrow samples were collected at the time of diagnosis, at day 15, at day 22 for patients with minimal residual disease (MRD) ≥1% on day 15, and at day 42 of induction (end of induction [EOI]) with peripheral blood count recovery per protocol. Peripheral blood samples (5ml in EDTA) were collected at diagnosis and days 8, 15, 22 and 42/EOI. Samples were centrifuged at 400g for 10 minutes and plasma was transferred to a separate tube. When cells were isolated, PBS was then added to the remaining sample in a 1 to 2 ratio, followed by density centrifugation at 400g for 30 minutes with Ficoll-Plus (Amersham) as per the manufacturer recommendations. Plasma and cells were then stored at −80 until DNA was extracted. Plasma separation was done within 6 hours from blood collection to limit cellular apoptosis and necrosis. CfDNA was extracted from 650μl – 1ml (average 950 μl) of plasma or non-cellular bone marrow (NCBM) plasma using Maxwell RSC ccfDNA Plasma Kit, according to the manufacturer’s protocol using 40μl elution buffer (Qiagen). CfDNA was quantified with a Qubit dsDNA High Sensitivity Kit. For each sample, all extracted cfDNA (i.e. without size selection) was input into NEBNext^®^ UltraII DNA Library Preparation Kit for Illumina (NEB #E7645) according to the manufacturer’s protocol except Kappa 10x primer mix was used for PCR enrichment of the adapter-ligated libraries and Qiagen EB buffer was used for elution. The traditional shearing step in library preparation was skipped to minimize contamination from non-apoptotic sources and increase the ctDNA purity of the library. Adapter combinations were selected per TruSeq DNA Sample Preparation Pooling Guidelines (Pooling Guidelines) for HiSeq.

**Figure 1.**
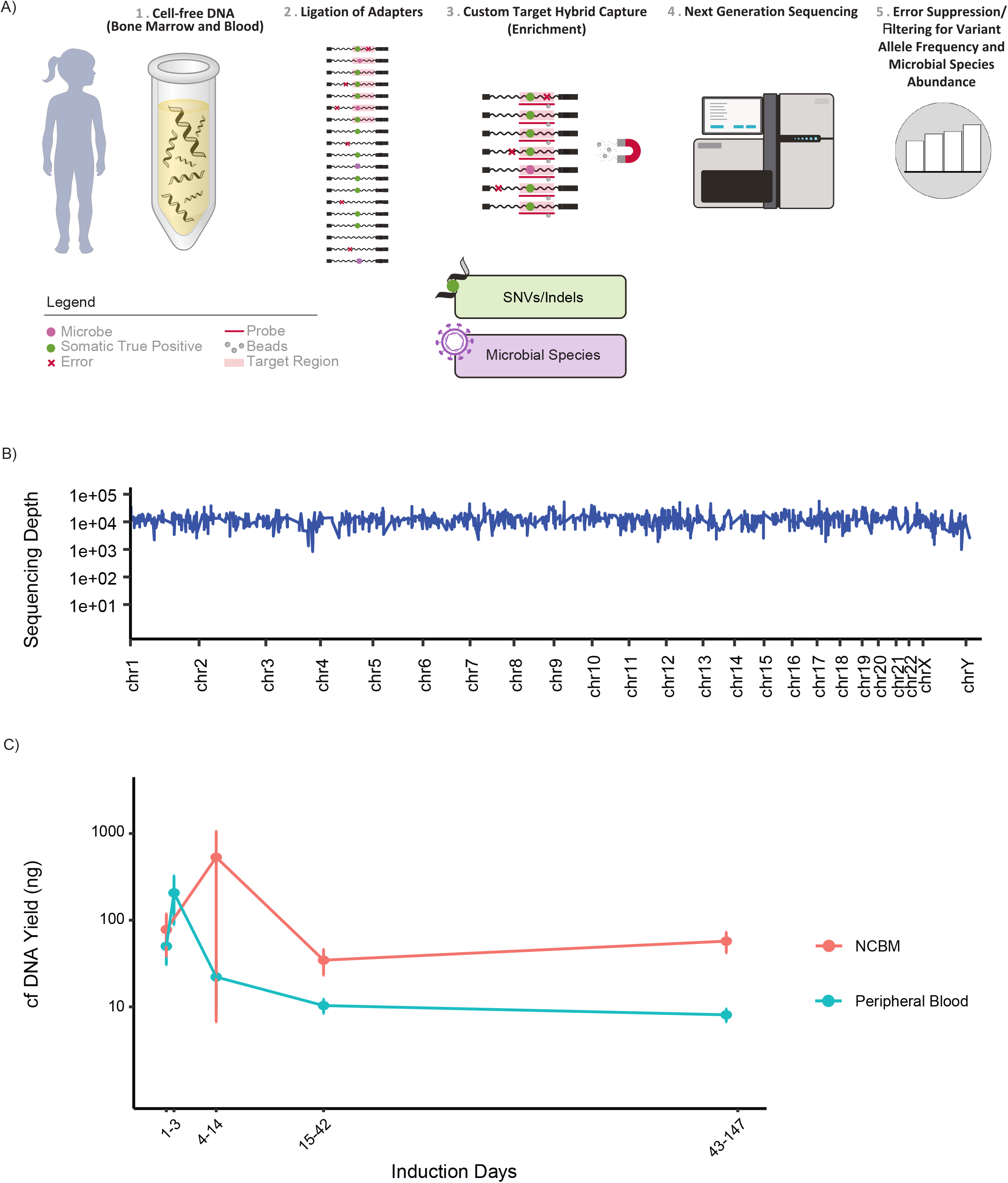
A) Workflow for contemporaneous and quantitative profiling of cancer variants and microbial species. Blood draw is followed by centrifugation for separation of cellular and acellular compartments. Probes spanning the target regions of the somatic leukemia panel as well as microbial species hybridize to targets and are enriched through magnetic separation followed by next generation sequencing. B) Depth of coverage is uniform across the panel, spanning all chromosomes. C) cfDNA yield from peripheral blood uptrends a few days after starting chemotherapy during tumor lysis whereas yield from non-cellular bone marrow (NCBM) increases during later phases of induction (while awaiting count recovery).

### Sequencing

Following library preparation and quantification, MiSeq sequencing was performed for assessment of the quality of the sequencing libraries prior to hybridization capture. For each case, libraries (equivalent to pre-defined time points) were pooled together in a 1:1 ratio into one capture. Whole exome sequencing (WES) was performed with an xGen Exome Research Panel (IDT) which consists of 429,826 probes spanning 39 Mb (19,396 genes) of the human genome covering 51 Mb of end-to-end tiled probe space. Sequencing was performed on the NextSeq instrument (Illumina).

### Custom Capture Panel Design

The design of our capture panel was informed by *a priori* sequencing of 600 ALL patients and the infectious capture panel was designed using probes specific to common human pathogens. For targeted hybridization-based capture of cfDNA, a fit-for-purpose gene panel covering 1668 exons (319.4kb) was used to supplement the xGen Pan-Cancer Panel (IDT) with previously defined somatic mutations in leukemia oncogene pathways, tumor suppressor genes, genes associated with relapse, genes associated with drug metabolism and glucocorticoid resistance, as well as genes identified from recent CRISPR/Cas9 screens and internal databases^39^. A deletion probe set was designed to regions surrounding heterozygous SNPs in genes commonly deleted in ALL. Heterozygous SNPs are from dbSNP (build 150) and found in ≥1% of samples. Only SNPs in non-repetitive regions with allele frequency of 0.3% to 0.5% were considered. Probes were 120bp in size as per the manufacturer specifications. Genomic coordinates for gene segments were retrieved from Halper-Stromberg et al^40^. Microbial probe sets were designed from molecular barcodes (16s/18s ribosomal RNA and ITS) and species-specific genes from human pathogens common in leukemia patients. Sequence data was derived from the NCBI gene database, OrthoDB, Virus Pathogen Database and Analysis Resource, Ribosomal Database Project, OrthoMCL Database, and EuPathDB databases.

### Variant Calling

For WES data analysis, FASTQ files were trimmed with Trimmomatic 0.35 to cut adapter and other manufacturer-specific sequences from reads. Alignment to the human reference genome GRCh38/hg38 was performed with a Burrows-Wheeler Aligner MEM 0.7.17. Duplicates were marked with Picard’s MarkDuplicates and local realignment and base-recalibration was achieved with GATK 4.1.4.1. Variant calling and filtering were performed following GATK’s best practices workflow for germline somatic pipeline for short variant discovery involving the HaplotypeCaller tool for calling and the VariantRecalibrator tool for filtering. Annotation using various databases was performed with ANNOVAR^41^.

For targeted sequencing using the customized ALL gene panel, NGS reads were de-multiplexed using Illumina’s bcl2fastq 2.20. Trimmomatic was again used for removing manufacturer-specific sequences and alignment performed using BWA MEM 0.7.17 and GRCh38/hg38. Analysis of this deep sequencing data was done using the error suppression tool CleanDeepSeq^42^.

### Microbial Filtering

The targeted sequencing data contains reads from both ALL panel and microbe panel. Using the targeted sequencing data, manufacturer-specific sequences were trimmed from the FASTQ files using Trimmomatic 0.35, then sequentially aligned to human reference genome GRCh38/hg38 and Macaca mulatta (Rhesus monkey) Mmul_10 reference genome using the stringent BWA ALN 0.7.17 algorithm. The remaining unaligned reads were then collected for further microbe analysis. Microbes from the unaligned reads were first identified using metagenomic classifier Kraken2 2.0.8, and reads supporting these microbes were put through the BLAST+ 2.10.0 algorithm for concordance check (Figure 3A)^43, 44^. False positive reads were identified as 1) fewer than 10 supporting reads from virus and 50 reads from bacteria and fungi, 2) less than 95% BLAST+ match on both forward and reverse reads, 3) forward reads and reverse reads do not match to the same kraken species, and 4) the contig length outside 120-600bp window.

**Figure 3.**
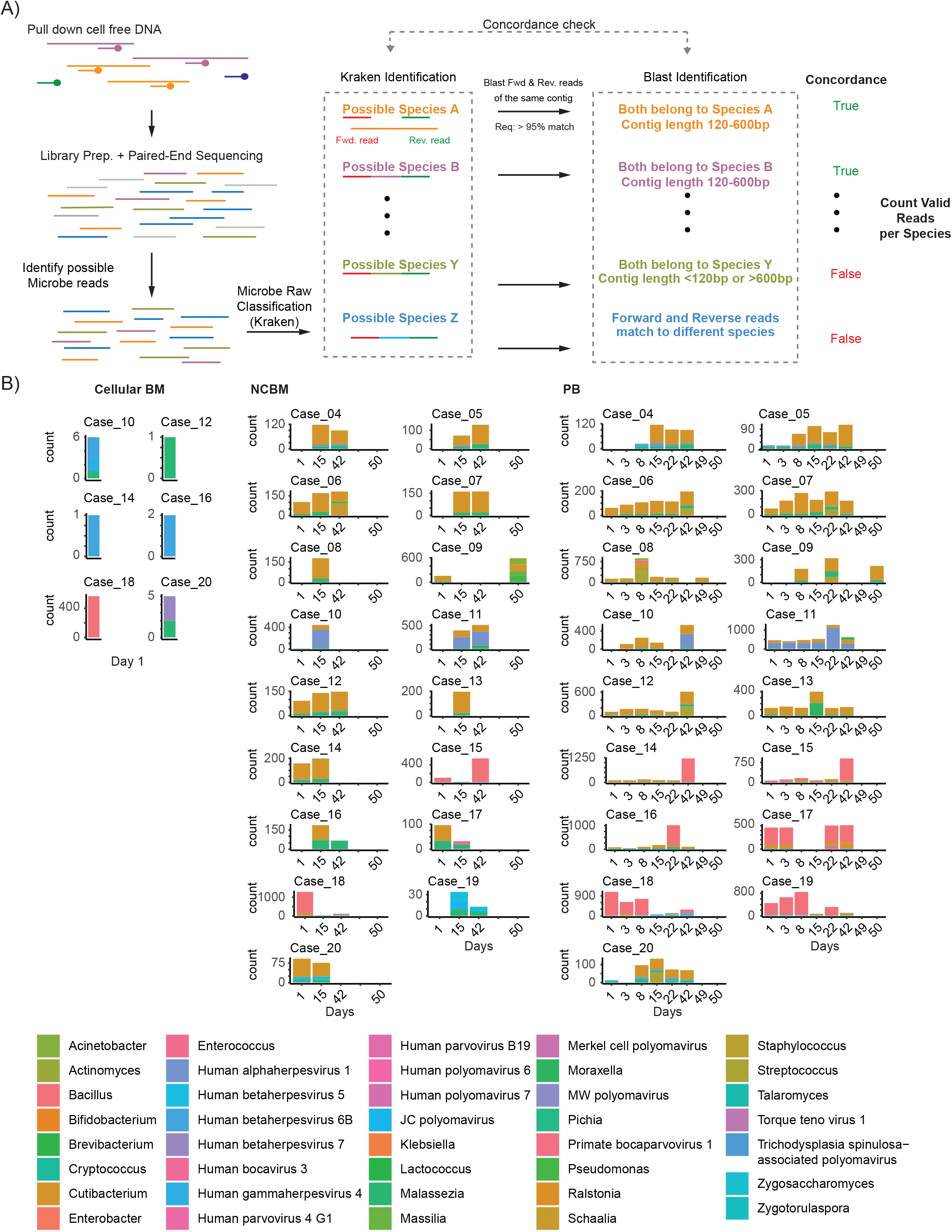
A) Workflow for identification of microbial species. Probes spanning the target regions of the microbial species hybridize to targets and are enriched through magnetic separation followed by next generation sequencing. Raw reads undergo concordance check through Kraken and Blast identification to determine valid number of reads per species. B) Microbial (viral, bacterial, and fungal) abundance in acellular bone marrow, peripheral blood, and cellular bone marrow (when detected) across induction days. Note axes for each case are fit to scale.

## Results

### Study Overview

168 samples underwent sequencing, prospectively collected from 20 newly diagnosed ALL patients over a 6-week course of induction therapy on the Total Therapy XVII clinical trial. Both cellular and plasma (cell-free) fractions were collected from peripheral blood and bone marrow to enable comparisons of distinct potential reservoirs of leukemia associated mutations and infectious microbial DNA. Panel coverage (Figure 1B) was found to be uniform over the target sequence with 100% of the target region covered at a mean depth of 12,884 reads.

At diagnosis, average plasma cfDNA yield was similar to NCBM cfDNA yield (mean 85 and 75 ng, respectively). Plasma cfDNA yield increased during the first few days of induction chemotherapy, likely a reflection of rapidly dying leukemic cells after beginning treatment. Similarly, NCBM cfDNA yield peaked at day 15 suggesting the cells within the bone marrow were still dying at a higher rate and at a later timepoint than the leukemia cells in peripheral blood cells. Thereafter, cfDNA concentration remained around 10ng per ml of blood throughout induction therapy whereas NCBM cfDNA yield increased in the latter part of induction therapy (Figure 1C), potentially reflecting the turnover of rejuvenating hematopoietic cells after induction therapy and during consolidation treatment.

### Patient Dynamics in Mutational VAF Clearance During Induction Therapy

The custom panel-based ctDNA NGS identified one or more variants at diagnosis in 17 of 20 patients (85%) (Figure 2A). Mutational clearance throughout induction chemotherapy was near universally observed. Each patient had an average of 2.3 detectable mutations (range 0-5, S.D. 1.3). 39 unique mutations were detected with only one instance of a shared mutation (KRAS p.G13D in cases 3 and 16). Distinct patterns of variant allele clearance were observed as patients underwent induction chemotherapy treatment. Generally, VAFs increased and then diminished throughout the course of induction therapy (Figure 2B) coincident with pharmacokinetics of chemotherapy and rapid cell death. Sequencing coverage was maintained over this time period (Figure 2C). Truncal mutations (defined as allele frequency >0.25 OR peripheral blood VAF >0.3 in both NCBM and peripheral blood) were found in 8 of 16 patients (71%) and included the genes NOTCH1, NRAS, KRAS, JAK2, NF1, and ADGRL2.

**Figure 2.**
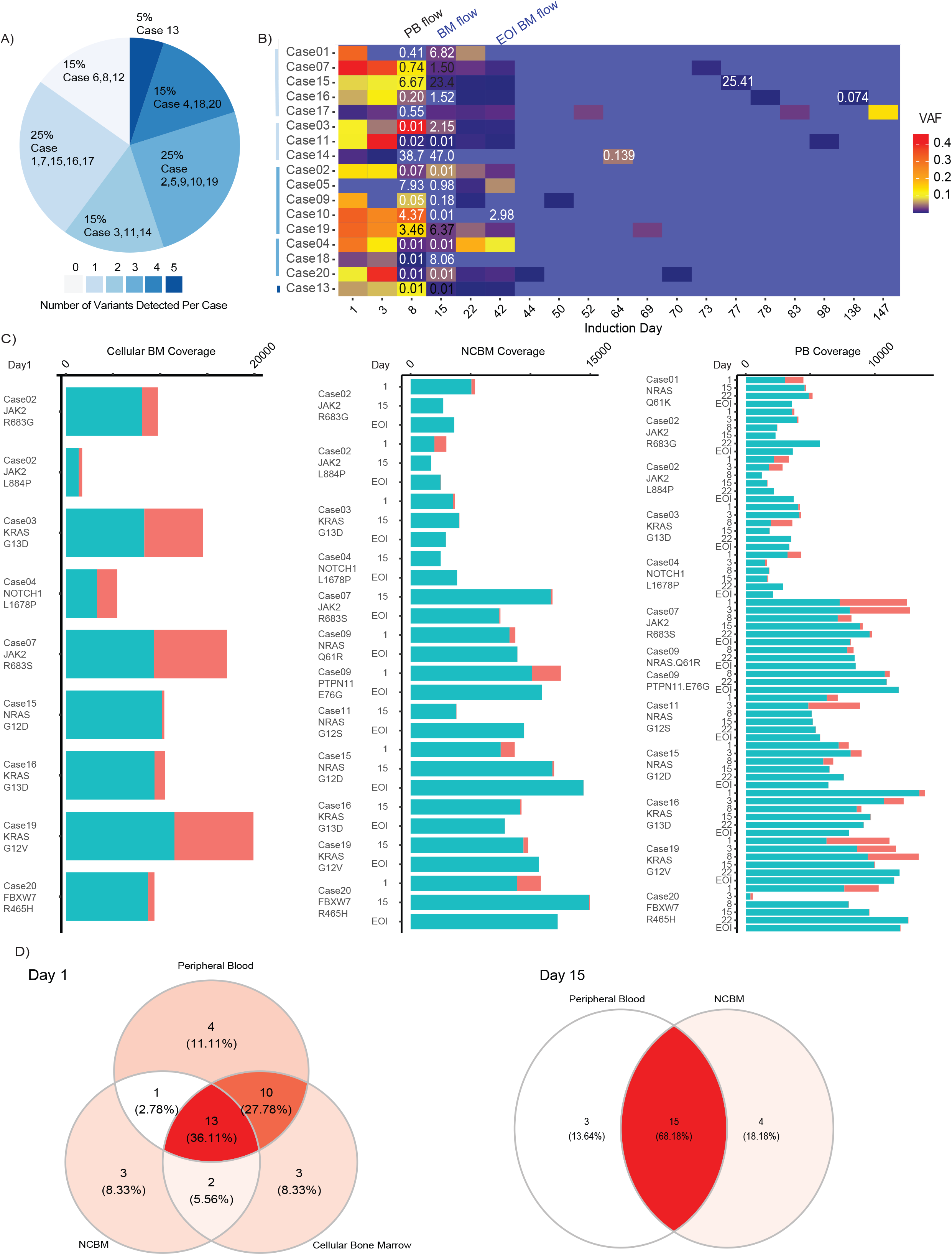
A) Number of unique variants detected per case. B) Mutational clearance as average VAF across all compartments throughout induction chemotherapy with flow cytometry MRD overlayed quantitatively for days 8 (from peripheral blood), 15 (from bone marrow) and 42 (from bone marrow). Mutational clearance was observed in most cases. In some instances (ex. Case 17 Day 150) mutations were detectable in the blood that were not observed in the bone marrow likely because of spatial heterogeneity. Variants (genes) were rarely shared across patients. Note Cases 10 and 14-16 MRD detectable by flow at end of induction. C) Coverage across cellular bone marrow, cell free bone marrow, and peripheral blood of selected variants known to be found in ALL patients. D) Correlations of variants detected on days 1 and 15 across compartments.

### Comparison of Cells, NCBM, PB cfDNA and Minimum Residual Disease

The concordance of variants detected across cellular, non-cellular bone marrow (NCBM) and peripheral blood cfDNA was assessed (Figure 2D). At diagnosis, peripheral blood VAFs correlated most strongly with VAFs detected in the cellular bone marrow (23/30 variants detected in both) with each compartment picking up a small number of unique variants (3 and 4 for cellular BM and peripheral blood cfDNA, respectively) suggesting that both compartments are capturing most of the same variant information (Supplemental Figure 1). Approximately a third of variants were simultaneously detected in all three compartments. Three cases showed VAFs >30% in the peripheral blood that were not detected in the bone marrow (Case 20: FBXW7, HUWE1, and MTUS2; Case 11: SMARCB4; Case 4: THSD7A). On Day 15, 68% of variants detected in the peripheral blood were also found in the cell free bone marrow. 4 patients (Cases 10, 14, 15, and 16) had detectable MRD by flow at EOI (Figure 2B). Several cases demonstrated detectable variants despite negative EOI MRD by flow. Conversely, one patient (Case 10) had persistent flow MRD at end of induction (day 42) without any detectable VAFs.

### Microbial Invasion Observed During Induction

We also established technical feasibility for detecting infectious microbes in both cells as well as cfDNA through our custom microbe panel and our novel microbe detection pipeline (Figure 3A). Of note, cases 1-3 did not undergo microbial enrichment as part of the workflow and did not have any microbes detected at any timepoints. In addition, we evaluated mcfDNA from 5 healthy controls and 5 patients with acute myelogenous leukemia (AML) at diagnosis as controls. Importantly, we found the reads assigned to a specific microbe spanned both the probe target regions and surrounding areas of the genome (Supplementary Figure 2).

In general, we found the changes in mcfDNA to be dynamic and the microbes detected to be recurrent across induction therapy within a patient (Figure 3B). When compared to healthy control samples, we found changes in the dominant microbe species detected in ALL patients. For example, bacterial species from the Ralstonia genus were detected in all healthy samples, but only one of the 126 ALL samples. Conversely, Cutibacterium were detected in most ALL samples but none of the healthy controls (Supplemental Figure 4), which may represent the ability of the commensal skin bacteria to invade the human host. Importantly, Bacillus species, a known cause of mortality of pediatric ALL patients^45^, were detected in a subset of patients. For fungal DNA, Malassezia species were detected in 60% of healthy controls and 13/17 (82%) of diagnostic ALL samples, but none of the AML samples (Supplementary Figure 5).

### Complex Virome Dynamics in ALL Patients During Induction Therapy

Trending of viral species detection during induction therapy in peripheral blood demonstrated reproducibility in species detected for each case (Figure 4 A) and compared to healthy controls as well as 5 age-matched AML cases. Similar to our bacterial analyses, all healthy control sample had a single viral species detected in all samples: Human herpesvirus 6 (HHV6B). In addition, only 1/17 (16%) of ALL diagnostic samples had HHV6B detected while 9/17 (53%) diagnostic samples had at least one viral species detected at diagnosis. 9/9 patients with a viral species detected at diagnosis had the same species detected in at least one subsequent sample, 8/9 had additional viral species detected over induction therapy, and 6/8 patients with no viral species detected at diagnosis subsequently had the detection of a DNA virus (Figure 3B). These results suggest that viral infection and reactivation are present at the time of diagnosis, but that the relative species composition changes over time as patients receive induction chemotherapy. To look for evidence that these viral species could be contributing to leukemia formation and persistence, we used the same enrichment strategy to examine the diagnostic bone marrow cellular fractions. We found a small number of sequencing reads from *cellular* bone marrow samples mapping to HHV6B in 3 patients and Human gammherpesvirus 4 (EBV) in one patient. However, those species were both found in normal healthy and AML samples, suggesting they are not uniquely required for the persistence of ALL cells at the time of diagnosis.

**Figure 4.**
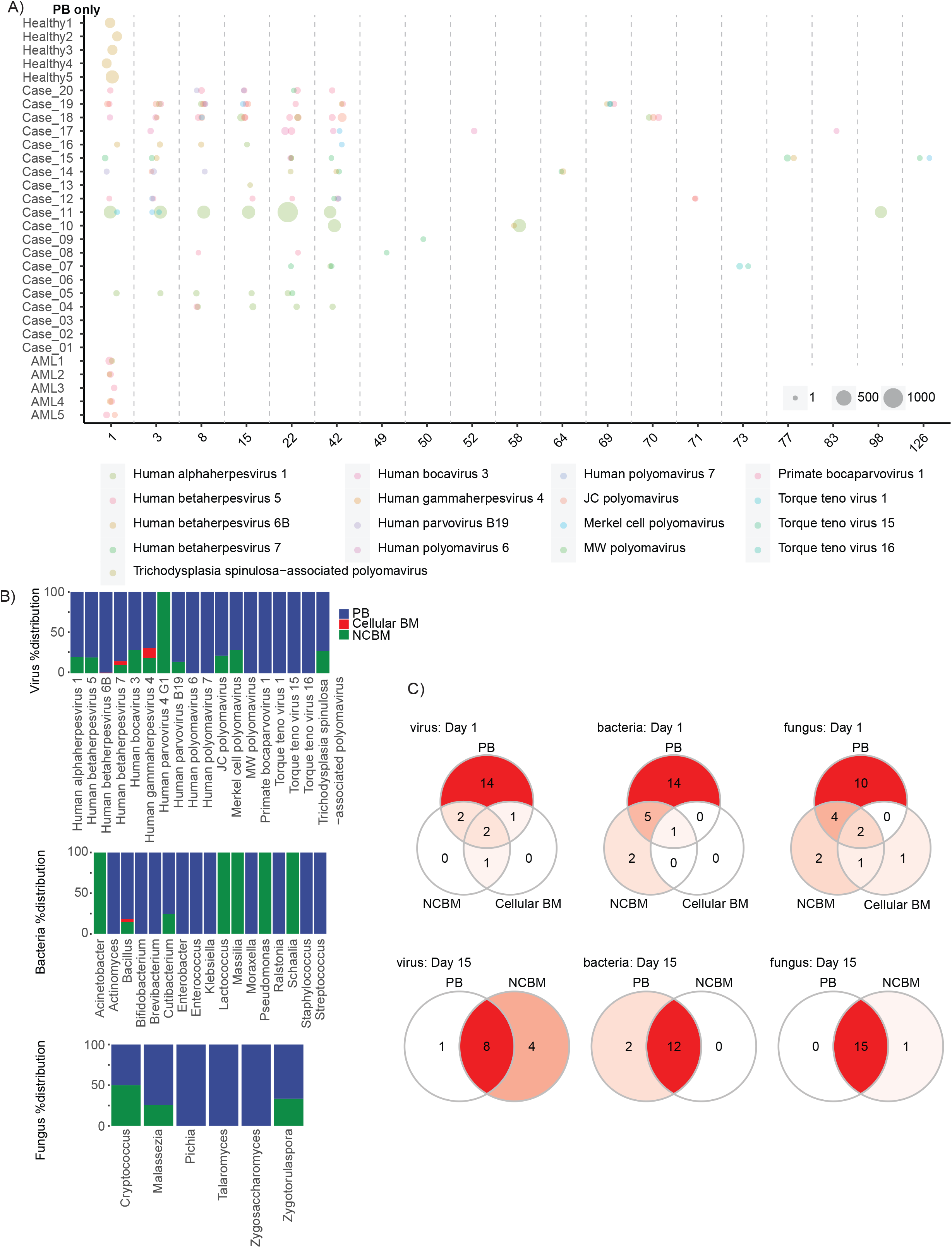
A) Peripheral blood viral species detection during induction therapy. Viral species were consistently detected for each case across induction days in peripheral blood. B) Representation of microbial species detected according to compartment across all cases. C) Concordance of fungal, viral, and bacterial species detected across compartments on Days 1 and 15.

The viral families detected in our ALL samples were Herpesviradae, Polyomaviridae, Parvoviridae, and Anelloviridae. Five different Herpesviradea were detected, and 16/17 patients had at least one viral species detected, suggesting Herpes virus reactivation is common in children undergoing induction therapy for ALL. Human alphaherpesvirus1 (HSV1) showed the most persistent detection in positive patients. Interestingly, none of these patients developed clinical symptoms that motivated clinicians to test them for HSV1 (Table 1). Polyomaviridae were the second most common family of viruses detected with 12/17 patients testing positive for at least one species while the Parvoviridae and Anneloviridae showed more sporadic detection.

### Common Features of Microbes in ALL Patients

We found several consistent microbe reservoir patterns within classes across patients. First, fungal and viral sequences tended to be found consistently in both the bone marrow and peripheral blood plasma while bacteria tended to be compartment-specific (Figure 4B). Interestingly, that pattern changes between day 1 and 15 when peripheral blood identified more species at day 1 whereas the two compartments are usually concordant on day 15 (Figure 4C). This suggests the organisms are in locations outside of the bone marrow sampling site at diagnosis and still detected in the peripheral blood, but that microbial DNA has become more widespread on day 15 (Supplemental Figure 6). Importantly, all 17 patients had negative viral and bacterial results when tested for clinical indications except case 10. This suggests that the microbes detected by cfDNA are below the clinical detection and/or sensitivity of current microbe diagnostics.

## Discussion

We have shown that a custom ctDNA capture strategy can be used to simultaneously follow disease persistence and microbe dynamics in patients with ALL. We generally found that PB ctDNA persistence generally correlates with disease persistence detected by flow cytometry, and that each compartment can detect disease when the other is negative. This suggests that the two approaches capture complementary information, with ctDNA having the added benefit of potentially identifying variants that are known to have qualitative differences in outcomes, such as NT5C2^29^ or PRSP1^46^ mutations resulting in thiopurine resistance and disease recurrence. Our comparisons of NCBM and PB plasma with the cellular leukemia cells at diagnosis found that PB and cellular leukemia cells most closely correlate at diagnosis, but that each compartment is able to capture distinct variants. After starting therapy, the NCBM and PB ctDNA were more closely correlated. These findings suggest that performing invasive bone marrow exams may not add much additional information for genomic testing. Additional studies on larger populations of patients are needed to further clarify these findings.

We also found evidence for dynamic changes in invasive microbes in ALL patients. We identified changes in the most common viral and bacterial species detected compared to healthy controls and found viral reactivation — especially of Herpesviridae and Polyomaviradae as patients underwent treatment. These microbes were all present at subclinical levels. Further work is needed to clarify whether they are altering the clinical course of these patients, including by directly causing infections or limiting the effectiveness of anticancer therapies by delaying administration of chemotherapy or altering the pharmacokinetics. We did not find direct evidence for active microbe infections contributing to the transformation or persistence of the leukemia cells. However, it is possible that one or more of these microbes could be contributing to the initial leukemic transformation but are dispensable as the malignant populations gain the ability to autonomously expand.

Our approach had several important limitations. First, we used a targeted approach using known mutations and microbes. Using a broader exome panel would enable us to capture unknown mutations that enable leukemic cell persistence through cancer evolution or iatrogenic mutagenesis, likely increasing the number of mutations detected per patient and sensitivity of our approach. Similarly, our microbe approach focused on known species. Performing a less biased experimental or computational approach, such as the depletion of human DNA and/or assembly of unmapped reads could uncover novel microbes. We acquired some information on the spatial dynamics of leukemic evolution and microbe invasion by comparing the bone marrow to the peripheral blood. Additional insights could be gained by accessing additional bone marrow sites and other tissue types in these patients.

## Conclusion

We have demonstrated that a customized hybrid capture NGS panel can noninvasively measure the two leading causes of mortality in pediatric ALL patients: leukemic disease burden as well as the invasion of infectious microbes. This proof-of-concept helps establish a technical and conceptual framework that we anticipate will be expanded and applied to different populations of patients with leukemias, as well as extended to additional cancers. This will help accelerate the attainment of biological insights into the temporal biology of host-microbe-leukemia interactions, including how those changes correlate with and alter anticancer therapy efficacy. We also anticipate fewer invasive bone marrow biopsies will be necessary as these methods improve with standardization and are validated for clinical use.

## Data Availability

The sequencing data from this study will be deposited into the Short Read Archive prior to publication.

## Acknowledgements

We would like to acknowledge Joshua Stokes from St. Jude Biomedical Communications for contributing to the figure generation. V.B. is an Anne T. and Robert M. Bass Endowed Fellow, supported by the Stanford Child Health Research Institute. C.G. is supported by a Career Award for Medical Scientists from the Burroughs Welcome Fund, NIH Director’s New Innovator Award (1DP2CA239145-01), and the Chan Zuckerberg Biohub. This work was also supported by St. Jude Cancer Center Developmental Funds (2P30CA021765-40), the Eleftheria Foundation, and ALSAC.

## Authorship Contributions

Conceptualization, HI, WE, CG

Methodology, VG, SY, YI, OM, CG

Investigation, VB, YX, DK, VG, RA, SY, YI, OM

Formal Analysis, YX, DK, VA, RA, OM, CG

Writing– Original Draft, VB, YX, CG

Writing– Review & Editing, VB, YX, CG

Visualization, VB, YX, DK, VG, VA, SY, OM

Funding Acquisition, WE, CG

Supervision, CP, HI, WE, CG

## References

1. Fan HC, Blumenfeld YJ, Chitkara U, Hudgins L, Quake SR. Noninvasive diagnosis of fetal aneuploidy by shotgun sequencing DNA from maternal blood. Proc Natl Acad Sci U S A. Oct 21 2008;105(42):16266-71. doi:10.1073/pnas.0808319105

2. Bianchi DW, Parker RL, Wentworth J, et al. DNA sequencing versus standard prenatal aneuploidy screening. N Engl J Med. Feb 27 2014;370(9):799-808. doi:10.1056/NEJMoa1311037

3. Diehl F, Schmidt K, Choti MA, et al. Circulating mutant DNA to assess tumor dynamics. Nat Med. Sep 2008;14(9):985-90. doi:10.1038/nm.1789

4. Forshew T, Murtaza M, Parkinson C, et al. Noninvasive identification and monitoring of cancer mutations by targeted deep sequencing of plasma DNA. Sci Transl Med. May 30 2012;4(136):136ra68. doi:10.1126/scitranslmed.3003726

5. Beaver JA, Jelovac D, Balukrishna S, et al. Detection of cancer DNA in plasma of patients with early-stage breast cancer. Clin Cancer Res. May 15 2014;20(10):2643-2650. doi:10.1158/1078-0432.CCR-13-2933

6. Newman AM, Bratman SV, To J, et al. An ultrasensitive method for quantitating circulating tumor DNA with broad patient coverage. Nat Med. May 2014;20(5):548-54. doi:10.1038/nm.3519

7. De Vlaminck I, Martin L, Kertesz M, et al. Noninvasive monitoring of infection and rejection after lung transplantation. Proc Natl Acad Sci U S A. Oct 27 2015;112(43):13336-41. doi:10.1073/pnas.1517494112

8. Blauwkamp TA, Thair S, Rosen MJ, et al. Analytical and clinical validation of a microbial cell-free DNA sequencing test for infectious disease. Nat Microbiol. Apr 2019;4(4):663-674. doi:10.1038/s41564-018-0349-6

9. Pui CH, Yang JJ, Hunger SP, et al. Childhood Acute Lymphoblastic Leukemia: Progress Through Collaboration. J Clin Oncol. Sep 20 2015;33(27):2938-48. doi:10.1200/JCO.2014.59.1636

10. O’Connor D, Bate J, Wade R, et al. Infection-related mortality in children with acute lymphoblastic leukemia: an analysis of infectious deaths on UKALL2003. Blood. Aug 14 2014;124(7):1056-61. doi:10.1182/blood-2014-03-560847

11. Mandel P, Metais P. Les acides nucléiques du plasma sanguin chez l’homme. C R Seances Soc Biol Fil. Feb 1948;142(3-4):241-3. Les acides nucleiques du plasma sanguin chez l’homme.

12. Leon SA, Shapiro B, Sklaroff DM, Yaros MJ. Free DNA in the serum of cancer patients and the effect of therapy. Cancer Res. Mar 1977;37(3):646-50.

13. Murtaza M, Dawson SJ, Tsui DW, et al. Non-invasive analysis of acquired resistance to cancer therapy by sequencing of plasma DNA. Nature. May 2 2013;497(7447):108-12. doi:10.1038/nature12065

14. Tie J, Wang Y, Tomasetti C, et al. Circulating tumor DNA analysis detects minimal residual disease and predicts recurrence in patients with stage II colon cancer. Sci Transl Med. Jul 6 2016;8(346):346ra92. doi:10.1126/scitranslmed.aaf6219

15. Provencio M, Torrente M, Calvo V, et al. Prognostic value of quantitative ctDNA levels in non small cell lung cancer patients. Oncotarget. Jan 2 2018;9(1):488-494. doi:10.18632/oncotarget.22470

16. Bidard FC, Madic J, Mariani P, et al. Detection rate and prognostic value of circulating tumor cells and circulating tumor DNA in metastatic uveal melanoma. Int J Cancer. Mar 1 2014;134(5):1207-13. doi:10.1002/ijc.28436

17. Wang Y, Li L, Cohen JD, et al. Prognostic Potential of Circulating Tumor DNA Measurement in Postoperative Surveillance of Nonmetastatic Colorectal Cancer. JAMA Oncol. May 9 2019;doi:10.1001/jamaoncol.2019.0512

18. Lennon AM, Buchanan AH, Kinde I, et al. Feasibility of blood testing combined with PET-CT to screen for cancer and guide intervention. Science. Jul 3 2020;369(6499)doi:10.1126/science.abb9601

19. Mattox AK, Bettegowda C, Zhou S, Papadopoulos N, Kinzler KW, Vogelstein B. Applications of liquid biopsies for cancer. Sci Transl Med. Aug 28 2019;11(507)doi:10.1126/scitranslmed.aay1984

20. Koh W, Pan W, Gawad C, et al. Noninvasive in vivo monitoring of tissue-specific global gene expression in humans. Proc Natl Acad Sci U S A. May 20 2014;111(20):7361-6. doi:10.1073/pnas.1405528111

21. Snyder MW, Kircher M, Hill AJ, Daza RM, Shendure J. Cell-free DNA Comprises an In Vivo Nucleosome Footprint that Informs Its Tissues-Of-Origin. Cell. Jan 14 2016;164(1-2):57-68. doi:10.1016/j.cell.2015.11.050

22. Kurtz DM, Scherer F, Jin MC, et al. Circulating Tumor DNA Measurements As Early Outcome Predictors in Diffuse Large B-Cell Lymphoma. J Clin Oncol. Oct 1 2018;36(28):2845-2853. doi:10.1200/JCO.2018.78.5246

23. Spina V, Bruscaggin A, Cuccaro A, et al. Circulating tumor DNA reveals genetics, clonal evolution, and residual disease in classical Hodgkin lymphoma. Blood. May 31 2018;131(22):2413-2425. doi:10.1182/blood-2017-11-812073

24. Desch AK, Hartung K, Botzen A, et al. Genotyping circulating tumor DNA of pediatric Hodgkin lymphoma. Leukemia. 01 2020;34(1):151-166. doi:10.1038/s41375-019-0541-6

25. Mullighan CG, Goorha S, Radtke I, et al. Genome-wide analysis of genetic alterations in acute lymphoblastic leukaemia. Nature. Apr 2007;446(7137):758-64. doi:10.1038/nature05690

26. Mullighan CG. Genomic profiling of B-progenitor acute lymphoblastic leukemia. Best Pract Res Clin Haematol. Dec 2011;24(4):489-503. doi:10.1016/j.beha.2011.09.004

27. Zhang J, Mullighan CG, Harvey RC, et al. Key pathways are frequently mutated in high-risk childhood acute lymphoblastic leukemia: a report from the Children’s Oncology Group. Blood. Sep 2011;118(11):3080-7. doi:10.1182/blood-2011-03-341412

28. Hogan LE, Meyer JA, Yang J, et al. Integrated genomic analysis of relapsed childhood acute lymphoblastic leukemia reveals therapeutic strategies. Blood. Nov 2011;118(19):5218-26. doi:10.1182/blood-2011-04-345595

29. Meyer JA, Wang J, Hogan LE, et al. Relapse-specific mutations in NT5C2 in childhood acute lymphoblastic leukemia. Nat Genet. Mar 2013;45(3):290-4. doi:10.1038/ng.2558

30. Ding LW, Sun QY, Tan KT, et al. Mutational Landscape of Pediatric Acute Lymphoblastic Leukemia. Cancer Res. 01 2017;77(2):390-400. doi:10.1158/0008-5472.CAN-16-1303

31. Ma X, Edmonson M, Yergeau D, et al. Rise and fall of subclones from diagnosis to relapse in pediatric B-acute lymphoblastic leukaemia. Nat Commun. Mar 2015;6:6604. doi:10.1038/ncomms7604

32. Wood B, Wu D, Crossley B, et al. Measurable residual disease detection by high-throughput sequencing improves risk stratification for pediatric B-ALL. Blood. 03 2018;131(12):1350-1359. doi:10.1182/blood-2017-09-806521

33. Andersson D, Fagman H, Dalin MG, Ståhlberg A. Circulating cell-free tumor DNA analysis in pediatric cancers. Mol Aspects Med. 04 2020;72:100819. doi:10.1016/j.mam.2019.09.003

34. De Vlaminck I, Khush KK, Strehl C, et al. Temporal response of the human virome to immunosuppression and antiviral therapy. Cell. Nov 21 2013;155(5):1178-87. doi:10.1016/j.cell.2013.10.034

35. Vu DL, Cordey S, Simonetta F, et al. Human pegivirus persistence in human blood virome after allogeneic haematopoietic stem-cell transplantation. Clin Microbiol Infect. Feb 2019;25(2):225-232. doi:10.1016/j.cmi.2018.05.004

36. Zanella MC, Cordey S, Kaiser L. Beyond Cytomegalovirus and Epstein-Barr Virus: a Review of Viruses Composing the Blood Virome of Solid Organ Transplant and Hematopoietic Stem Cell Transplant Recipients. Clin Microbiol Rev. Sep 16 2020;33(4)doi:10.1128/CMR.00027-20

37. Wilson MR, Sample HA, Zorn KC, et al. Clinical Metagenomic Sequencing for Diagnosis of Meningitis and Encephalitis. N Engl J Med. Jun 13 2019;380(24):2327-2340. doi:10.1056/NEJMoa1803396

38. Goggin KP, Gonzalez-Pena V, Inaba Y, et al. Evaluation of Plasma Microbial Cell-Free DNA Sequencing to Predict Bloodstream Infection in Pediatric Patients With Relapsed or Refractory Cancer. JAMA Oncol. Apr 1 2020;6(4):552-556. doi:10.1001/jamaoncol.2019.4120

39. Autry RJ, Paugh SW, Carter R, et al. Integrative genomic analyses reveal mechanisms of glucocorticoid resistance in acute lymphoblastic leukemia. Nat Cancer. Mar 2020;1(3):329-344. doi:10.1038/s43018-020-0037-3

40. Halper-Stromberg E, Steranka J, Giraldo-Castillo N, Fuller T, Desiderio S, Burns KH. Fine mapping of V(D)J recombinase mediated rearrangements in human lymphoid malignancies. BMC Genomics. Aug 2013;14:565. doi:10.1186/1471-2164-14-565

41. Wang K, Li M, Hakonarson H. ANNOVAR: functional annotation of genetic variants from high-throughput sequencing data. Nucleic Acids Res. Sep 2010;38(16):e164. doi:10.1093/nar/gkq603

42. Ma X, Shao Y, Tian L, et al. Analysis of error profiles in deep next-generation sequencing data. Genome Biol. Mar 14 2019;20(1):50. doi:10.1186/s13059-019-1659-6

43. Wood DE, Lu J, Langmead B. Improved metagenomic analysis with Kraken 2. Genome Biol. 11 2019;20(1):257. doi:10.1186/s13059-019-1891-0

44. Camacho C, Coulouris G, Avagyan V, et al. BLAST+: architecture and applications. BMC Bioinformatics. Dec 2009;10:421. doi:10.1186/1471-2105-10-421

45. Chou YL, Cheng SN, Hsieh KH, Wang CC, Chen SJ, Lo WT. Bacillus cereus septicemia in a patient with acute lymphoblastic leukemia: A case report and review of the literature. J Microbiol Immunol Infect. Jun 2016;49(3):448-51. doi:10.1016/j.jmii.2013.06.010

46. Li B, Li H, Bai Y, et al. Negative feedback-defective PRPS1 mutants drive thiopurine resistance in relapsed childhood ALL. Nat Med. Jun 2015;21(6):563-71. doi:10.1038/nm.3840

