## Supplementary material for "Simultaneous Monitoring of Disease and Microbe Dynamics Through Plasma DNA Sequencing in Pediatric Patients with Acute Lymphoblastic Leukemia": Table 1

| Case | Infectious Workup During Induction Chemotherapy |
| --- | --- |
| 1 | Blood cultures negative, EBV capsid IgG positive, VZV IgG positive, bartonella/toxo/CMV negative |
| 2 | Blood cultures negative, EBV nuclear and capsid IgG positive, bartonella/toxo negative (CMV/VZV negative) |
| 3 | Blood and stool cultures negative, histoplasma, norovirus, and C-diff negative |
| 4 | EBV nuclear and capsid IgG positive, CMV IgG positive, VZV IgG positive |
| 5 | Blood culture negative |
| 6 | Rectal surveillance culture positive, respiratory panel negative, blood cultures negative |
| 7 | Blood culture negative, respiratory panel negative |
| 8 | Urine culture positive, blood cultures negative, EBV nuclear and capsid IgG positive, CMV IgG positive, VZV IgG positive |
| 9 | Negative blood cultures, EBV nuclear and capsid IgG positive |
| 10 | Positive blood cultures NOS, CSF infectious PCRs negative |
| 11 | EBV negative, VZV IgG pos |
| 12 | Pan culture negative, EBV nuclear and capsid IgG positive, CMV IgG positive, VZV IgG positive, Histoplasma antibody yeast antigen high (1:8) but resulted as none detected |
| 13 | Multiple blood cultures negative, EBV nuclear and capsid IgG positive, VZV IgG positive |
| 14 | Pan culture negative |
| 15 | Blood culture negative |
| 16 | EBV capsid IgG pos, VZV IgG pos, respiratory panel negative |
| 17 | EBV negative, VZV IgG pos |
| 18 | Blood culture negative, C Diff Toxin PCR negative |
| 19 | Blood culture negative |
| 20 | N/A |
