## Supplementary material for "Simultaneous Monitoring of Disease and Microbe Dynamics Through Plasma DNA Sequencing in Pediatric Patients with Acute Lymphoblastic Leukemia": Table and Supplementary Figure Legends

Table 1. Summary of infectious disease clinical tests ordered during induction chemotherapy. Blood cultures ordered during episodes of fever or other suspicion of infection.

--

Supplementary Figure 1. Correlation of variants detected in non-cellular bone marrow (NCBM) with peripheral blood and cellular bone marrow at days 1 and 15 for A) all VAFs and B) VAFs less than 10%.

Supplementary Figure 2. Integrative Genomics Viewer (IGV) illustrating microbial sample sequencing reads assigned to a specific microbe span both the probe target regions and surrounding areas of the genome.

Supplementary Figure 3. Quantitative trends of viral species detected throughout induction chemotherapy in non-cellular bone marrow (NCBM) and cellular bone marrow.

Supplementary Figure 4. Quantitative trends of bacterial species detected throughout induction chemotherapy in peripheral blood, non-cellular bone marrow (NCBM), and cellular bone marrow.

Supplementary Figure 5. Quantitative trends of bacterial species detected throughout induction chemotherapy in peripheral blood, non-cellular bone marrow (NCBM), and cellular bone marrow.

Supplementary Figure 6. Correlation of microbial species detected between non-cellular bone marrow (NCBM) and peripheral blood at days 1, 15, and 42.
