## Supplementary Figures for "Simultaneous Monitoring of Disease and Microbe Dynamics Through Plasma DNA Sequencing in Pediatric Patients with Acute Lymphoblastic Leukemia"

**Supplemental Figure 1.**

A)

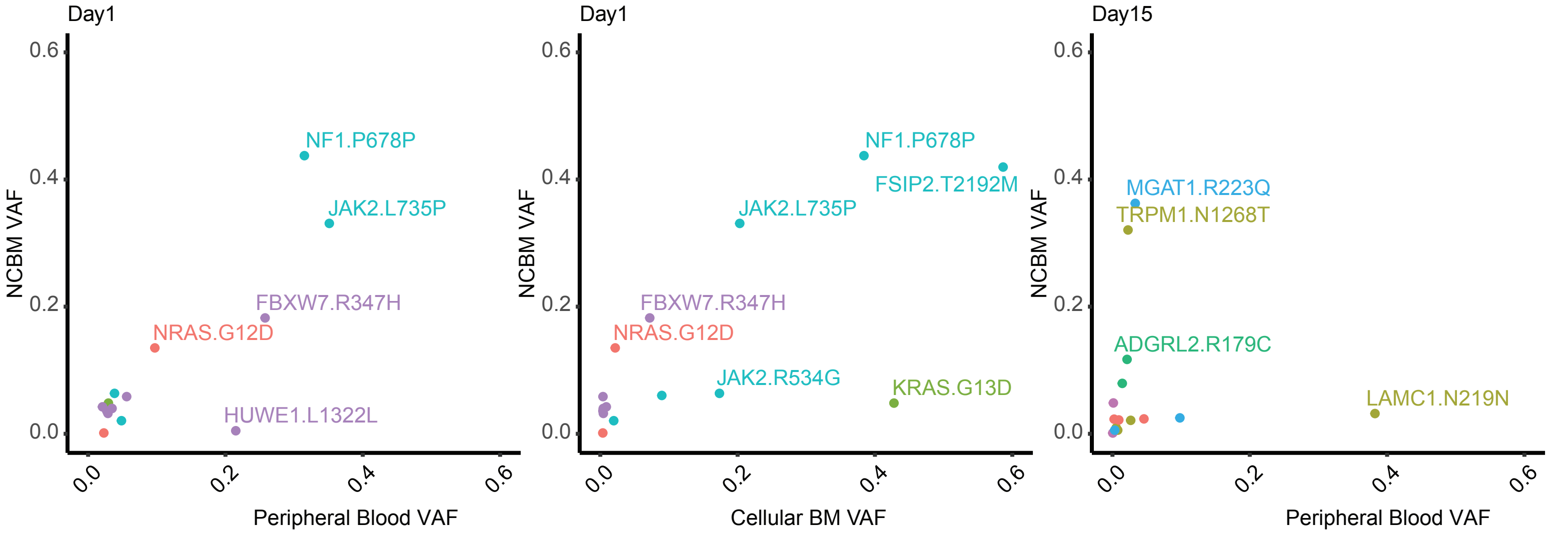


B)

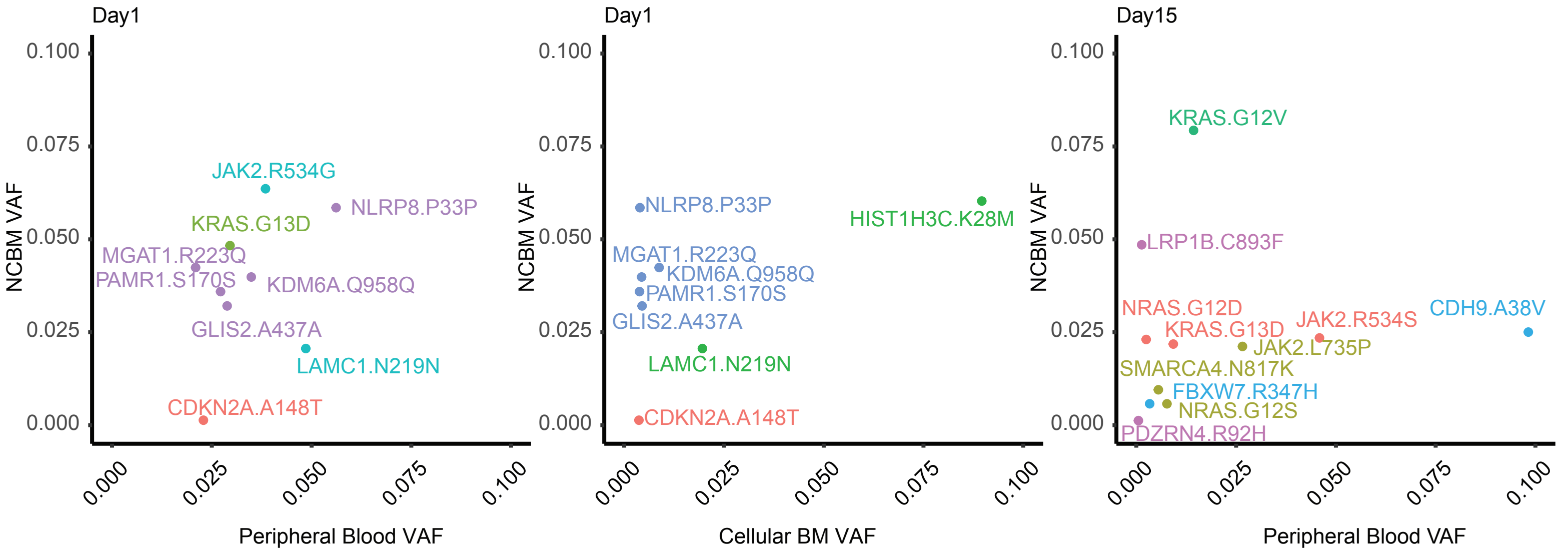

**Supplemental Figure 2.**

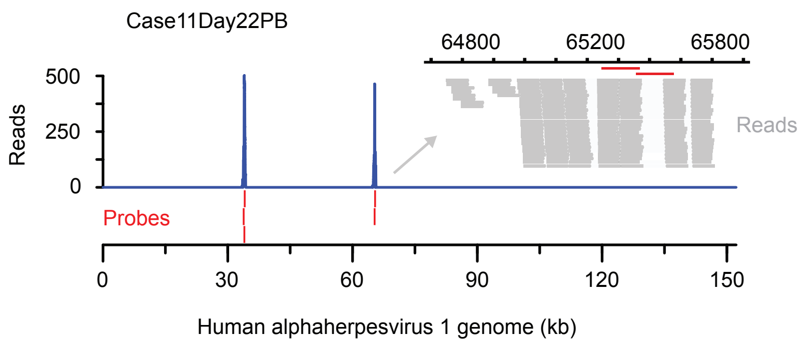


**Supplemental Figure 3.**

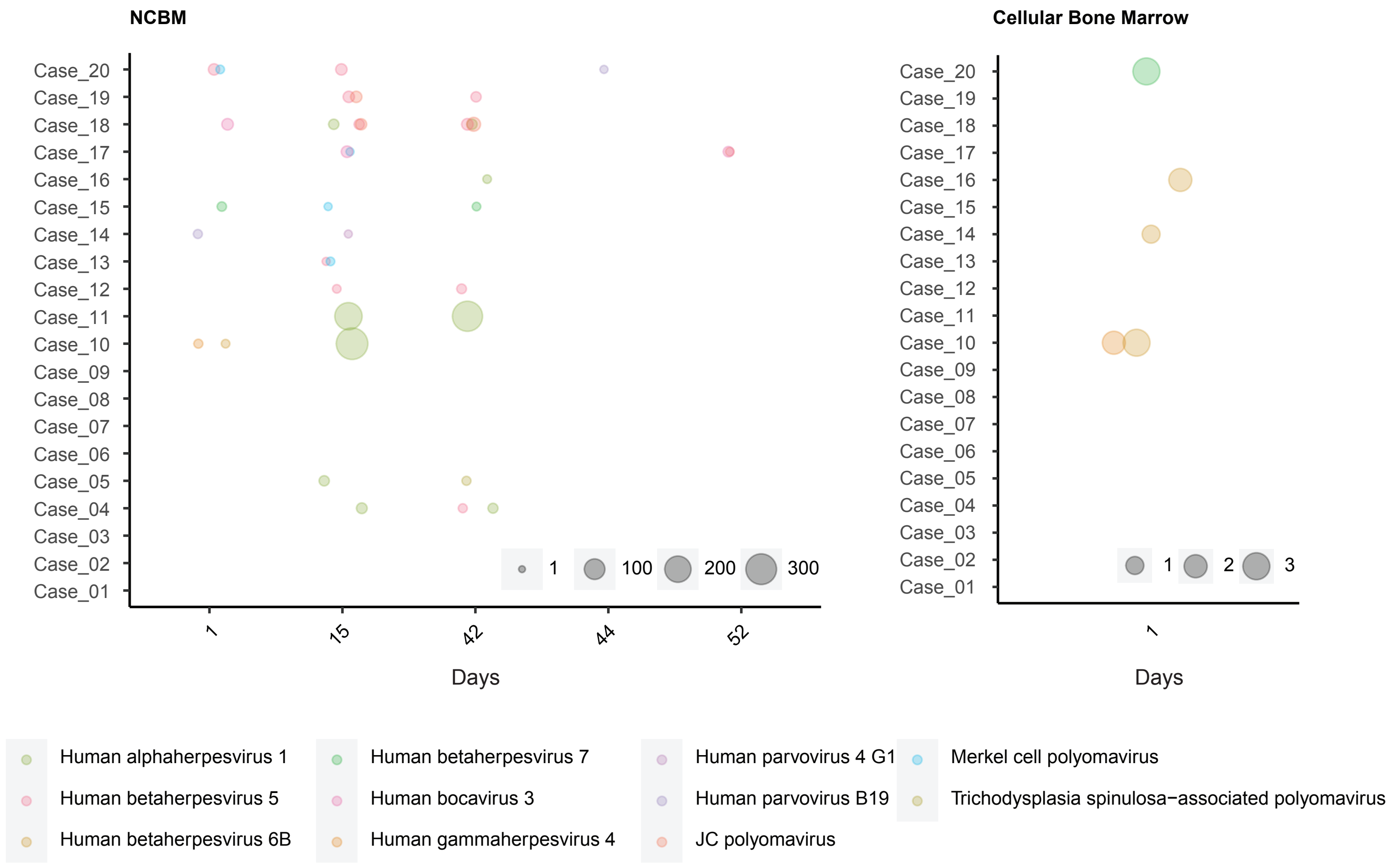

**Supplemental Figure 4.**

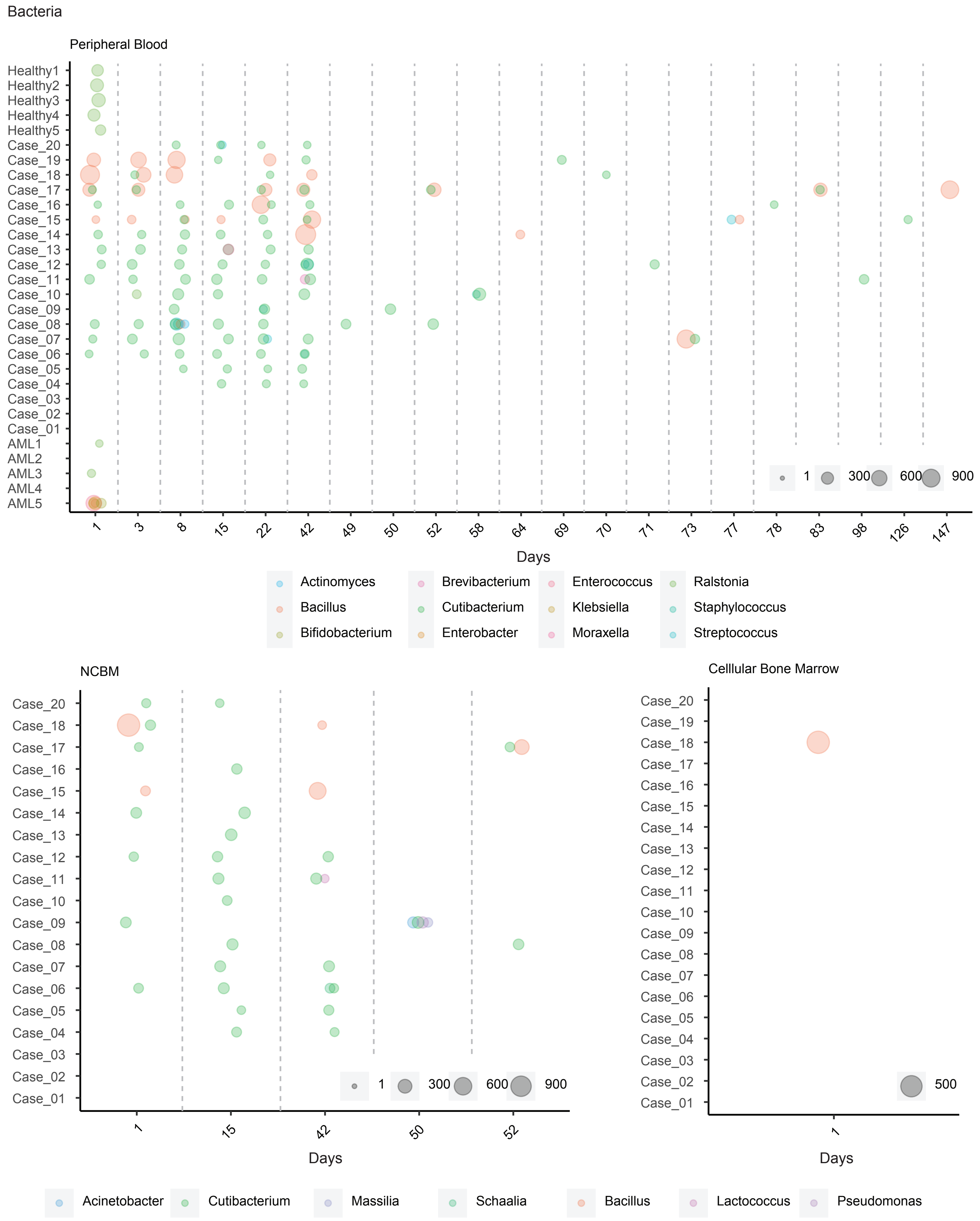

**Supplemental Figure 5.**

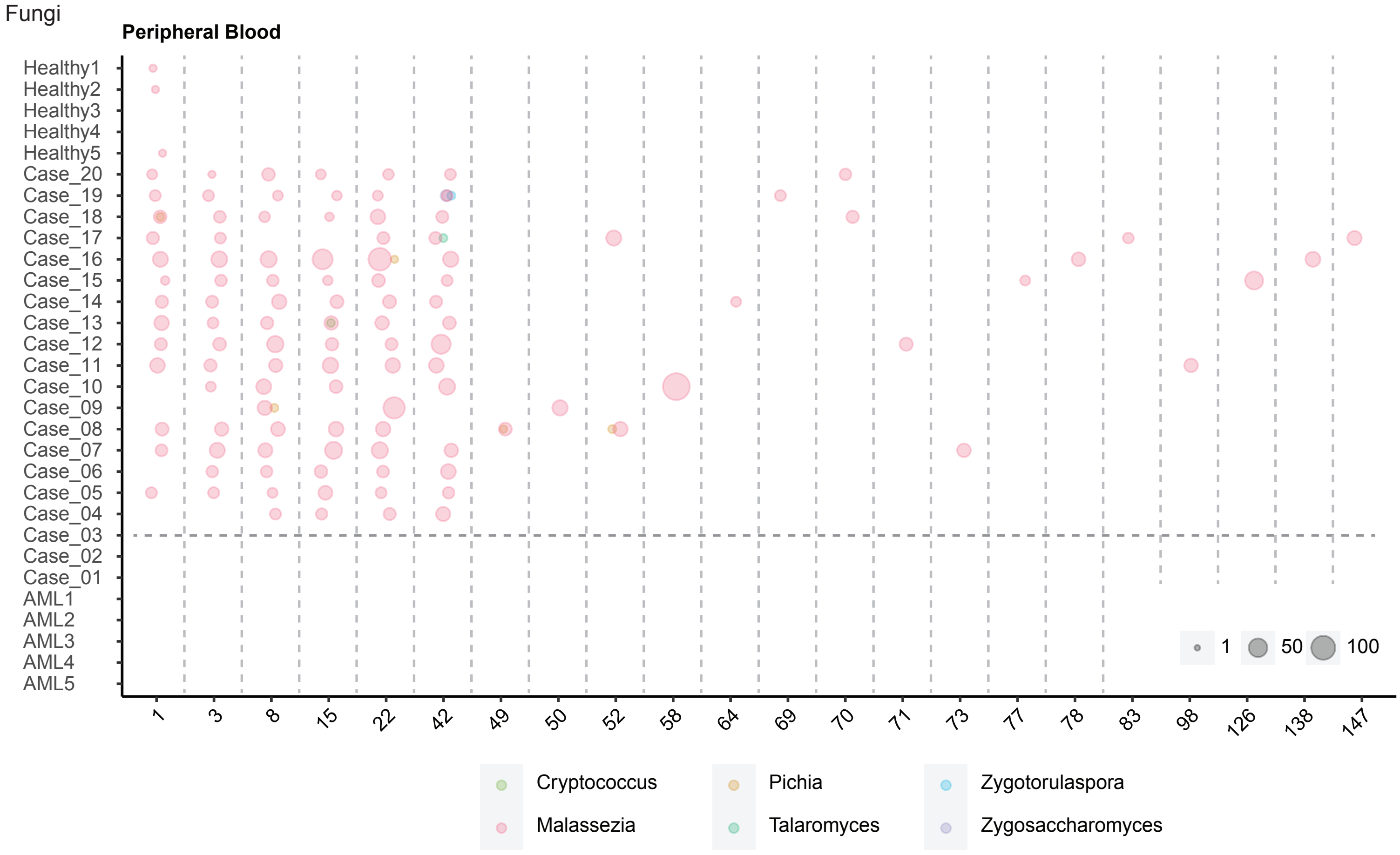


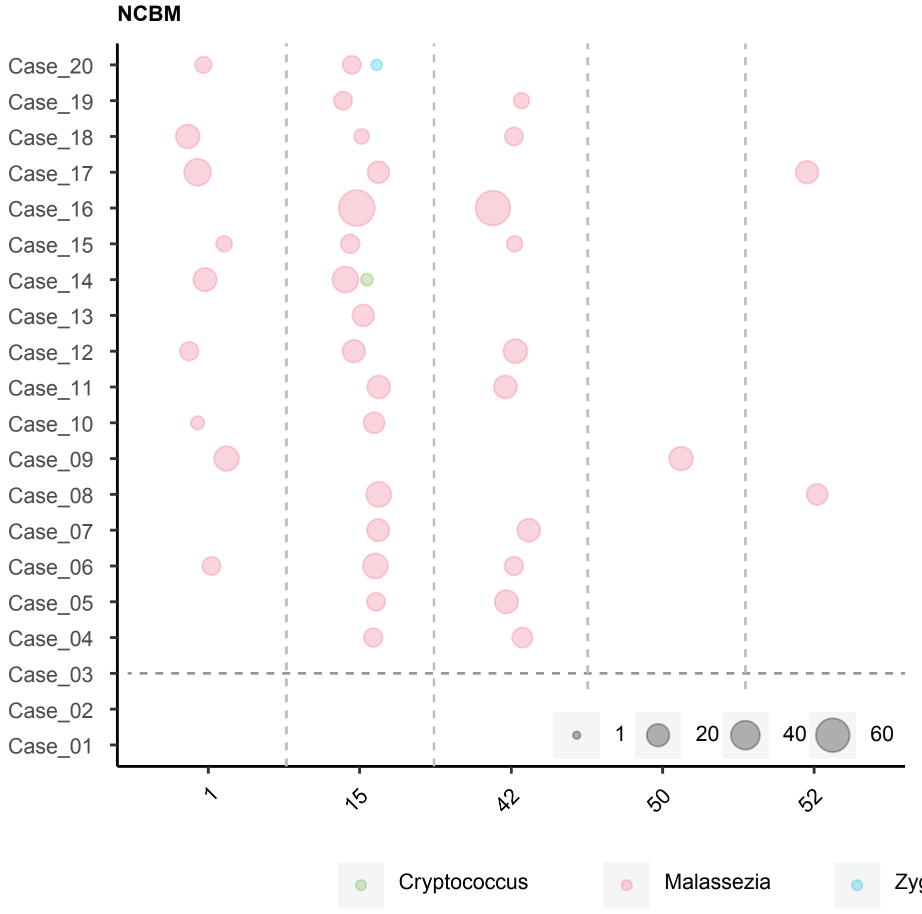


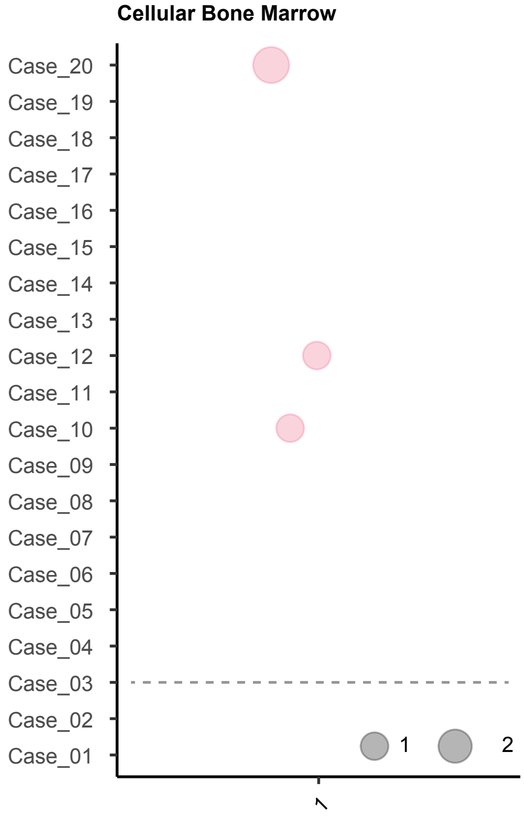

**Supplemental Figure 6.**

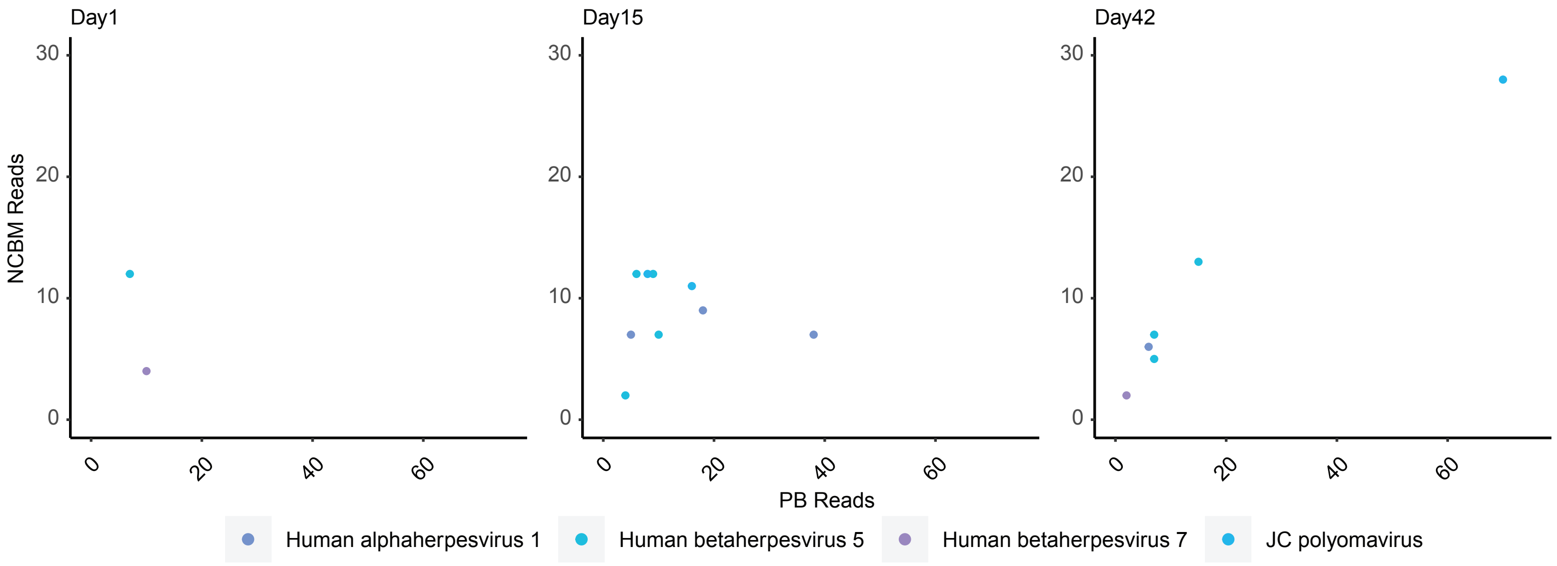


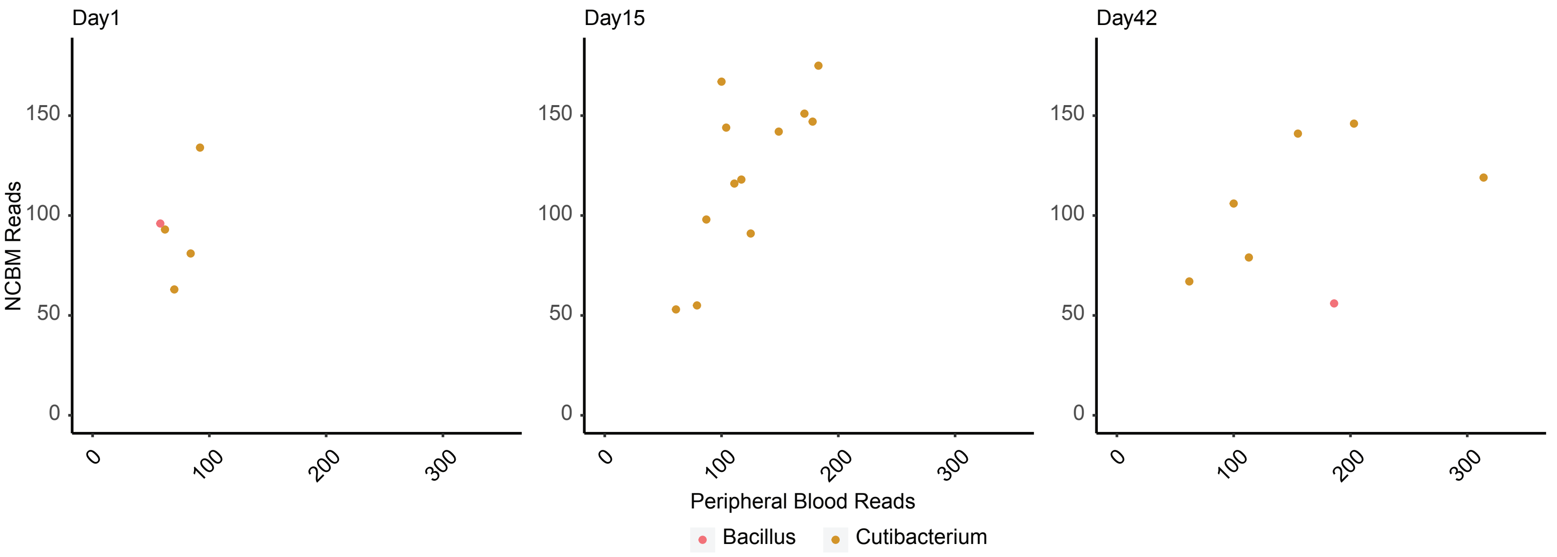


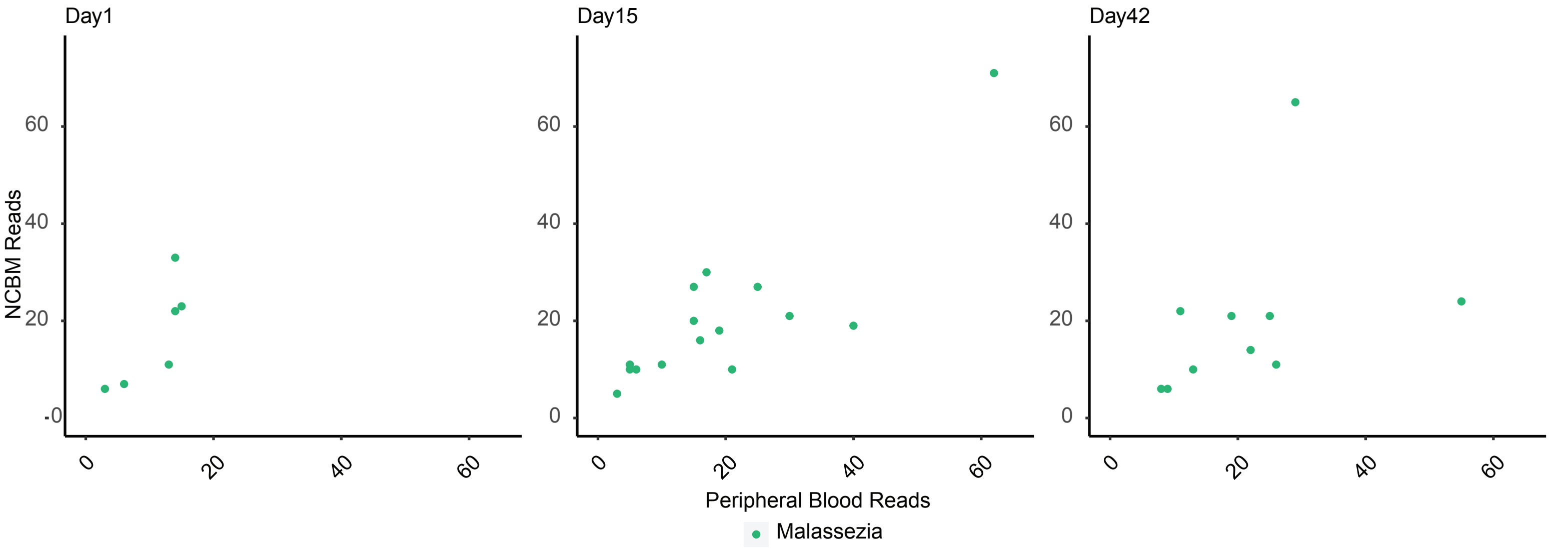
